# Effectiveness of casirivimab and imdevimab, and sotrovimab during Delta variant surge: a prospective cohort study and comparative effectiveness randomized trial

**DOI:** 10.1101/2021.12.23.21268244

**Authors:** David T. Huang, Erin K. McCreary, J. Ryan Bariola, Tami E. Minnier, Richard J. Wadas, Judith A. Shovel, Debbie Albin, Oscar C. Marroquin, Kevin E. Kip, Kevin Collins, Mark Schmidhofer, Mary Kay Wisniewski, David A. Nace, Colleen Sullivan, Meredith Axe, Russell Meyers, Alexandra Weissman, William Garrard, Octavia M. Peck-Palmer, Alan Wells, Robert D. Bart, Anne Yang, Lindsay R. Berry, Scott Berry, Amy M. Crawford, Anna McGlothlin, Tina Khadem, Kelsey Linstrum, Stephanie K. Montgomery, Daniel Ricketts, Jason N. Kennedy, Caroline J. Pidro, Rachel L. Zapf, Paula L. Kip, Ghady Haidar, Graham M. Snyder, Bryan J. McVerry, Donald M. Yealy, Derek C. Angus, Christopher W. Seymour

## Abstract

**IMPORTANCE:** The effectiveness of monoclonal antibodies (mAbs), casirivimab and imdevimab, and sotrovimab, for patients with mild to moderate COVID-19 from the Delta variant is unknown.

**OBJECTIVE:** To evaluate the effectiveness of mAbs for the Delta variant compared to no treatment, and the comparative effectiveness between mAbs.

**DESIGN, SETTING, AND PARTICIPANTS:** Two parallel studies among patients who met Emergency Use Authorization criteria for mAbs from July 14, 2021 to September 29, 2021: i.) prospective observational cohort study comparing mAb treatment to no mAb treatment and, ii.) Bayesian adaptive randomized trial comparing the effectiveness of casirivimab-imdevimab versus sotrovimab. In the observational study, we compared eligible patients who received mAb at an outpatient infusion center at UPMC, to nontreated patients with a positive SARS-CoV-2 test. In the comparative effectiveness trial, we randomly allocated casirivimab-imdevimab or sotrovimab to patients presenting to infusion centers and emergency departments, per system therapeutic interchange policy.

**EXPOSURE:** Intravenous mAb per their EUA criteria.

**MAIN OUTCOMES AND MEASURES:** For the observational study, risk ratio estimates for hospitalization or death by 28 days were compared between mAb treatment to no mAb treatment using propensity matched models. For the comparative effectiveness trial, the primary outcome was hospital-free days (days alive and free of hospital) within 28 days, where patients who died were assigned -1 day) in a Bayesian cumulative logistic model, adjusted for treatment location, age, sex, and time. Inferiority was defined as a 99% posterior probability of an odds ratio <1. Equivalence was defined as a 95% posterior probability that the odds ratio is within a given bound.

**RESULTS:** Among 3,558 patients receiving mAb, the mean age was 54 (SD 18 years), 1,511 (43%) were treated in an infusion center, and 450 (13%) were hospitalized or died by day 28. In propensity matched models, mAb treatment was associated with reduced risk of hospitalization or death compared to no treatment (risk ratio (RR)=0.40, 95% CI: 0.28–0.57). Both casirivimab and imdevimab (RR=0.31, 95% CI: 0.20–0.50), and sotrovimab (RR=0.60, 95% CI: 0.37–1.00) reduced hospitalization or death compared to no mAb treatment. Among patients allocated randomly to casirivimab and imdevimab (n=2,454) or sotrovimab (n=1,104), the median hospital-free days were 28 (IQR 28–28) for both groups, 28-day mortality was 0.5% (n=12) and 0.6% (n=7), and hospitalization by day 28 was 12% (n=291) and 12% (n=140), respectively. Compared to casirivimab and imdevimab, the median adjusted odds ratio for hospital-free days was 0.88 (95% credible interval, 0.70–1.11) for sotrovimab. This odds ratio yielded 86% probability of inferiority of sotrovimab versus casirivimab and imdevimab, and 79% probability of equivalence.

**CONCLUSIONS AND RELEVANCE:** In non-hospitalized patients with mild to moderate COVID-19 due to the Delta variant, casirivimab and imdevimab and sotrovimab were both associated with a reduced risk of hospitalization or death. The comparative effectiveness of mAbs appeared similar, though prespecified criteria for statistical inferiority or equivalence were not met.

**TRIAL REGISTRATION:** ClinicalTrials.gov: NCT04790786

**Key Points:** *Question:* In non-hospitalized patients with mild to moderate COVID-19 due to the Delta variant, what is the effectiveness of monoclonal antibodies (mAb) compared to no treatment, and what is the comparative effectiveness between mAb?

*Findings:* Among 3,069 patients, mAb treatment (casirivimab and imdevimab or sotrovimab) was associated with reduced risk of hospitalization or death by 28 days compared to no treatment (risk ratio=0.40, 95% CI: 0.28–0.57). In a Bayesian randomized comparative effectiveness trial of casirivimab and imdevimab vs. sotrovimab in 3,558 patients, the median hospital–free days were 28 days for both groups. Compared to casirivimab-imdevimab, the median adjusted odds ratio for hospital-free days was 0.88 (95% credible interval, 0.70–1.11) for sotrovimab, an 86% probability of inferiority of sotrovimab versus casirivimab and imdevimab, and 79% probability of equivalence.

*Meaning:* In non-hospitalized patients with mild to moderate COVID-19 due to the Delta variant, casirivimab and imdevimab and sotrovimab were associated with reduced risk of hospitalization or death compared to no treatment. The comparative effectiveness of mAbs appeared similar, though prespecified criteria for statistical inferiority or equivalence were not met.

## Introduction

Monoclonal antibodies (mAbs) were granted U.S. Food and Drug Administration (FDA) Emergency Use Authorization (EUA) for treatment of mild to moderate COVID-19.^1, 2^ However, their effectiveness with the Delta variant is unknown.

In February 2021, UPMC partnered with the U.S. Federal COVID-19 Response Team and received bamlanivimab, bamlanivimab-etesevimab, and casirivimab-imdevimab to expand clinical use and evaluate their effectiveness using a learning health system approach. This approach embeds knowledge generation into daily practice to seek continuous improvement in care. ^3, 4, 5^ In April 2021, the EUA for bamlanivimab was revoked, and in June 2021, UPMC partnered with GlaxoSmithKline and Vir Biotechnology to make sotrovimab available to EUA-eligible patients and evaluate its effectiveness. Because the U.S. government paused distribution of bamlanivimab-etesevimab, and the Delta variant became dominant in the U.S in summer 2021, we evaluated the effectiveness of casirivimab-imdevimab and sotrovimab from July 2021 to September 2021.^6,7^

This report presents an evaluation of casirivimab-imdevimab and sotrovimab from July 14, 2021 to September 29, 2021, with results released due to the Delta variant public health crisis. This study sought to determine, i) the effectiveness of mAbs for the Delta variant compared to no treatment, and ii) the comparative effectiveness between mAbs.

## Methods

This report includes two studies: i) a propensity score matched observational study of mAb treatment versus no mAb treatment (UPMC Quality Improvement Review Committee Project ID 2882 and Project ID 3116), and ii) a randomized comparative effectiveness trial of casirivimab and imdevimab versus sotrovimab (UPMC Quality Improvement Committee Project ID 3280 and University of Pittsburgh Institutional Review Board (IRB) STUDY21020179).

### Study setting and data sources

UPMC is a 40-hospital integrated health system principally in central and western Pennsylvania. After an EUA was granted for bamlanivimab on November 9, 2020, ^8^ UPMC developed a mAb treatment infrastructure.^9, 10^ On November 21, 2020, the UPMC Pharmacy and Therapeutics Committee wrote a therapeutic interchange policy in response to the issuance of an EUA for casirivimab and imdevimab and unpredictable mAb supply. The policy considered all available mAbs equivalent; a patient could receive any mAb based on local inventory. The policy was updated to include sotrovimab on July 9, 2021. All pharmacies supplying all infusion sites had equal opportunity to order any available mAb from a central supply facility. Prescribers were required to provide and review all mAb EUA Fact Sheets with the patient at time of referral and explain the patient could receive any EUA-governed mAb under the therapeutic interchange policy. EUA-eligible patients were referred via the electronic medical record (EMR) systems for UPMC providers and by paper order for non-UPMC prescribers. A centralized team with nurses, administrators, pharmacists, and physicians reviewed orders daily to confirm criteria for receipt. Decentralized nursing teams then contacted and scheduled eligible patients for infusions.

We used the EMR to access all key clinical data including detailed sociodemographic and medical history data, and hospitalization after mAb treatment, and augmented by manual review and data collection. We linked the deidentified primary data sources using common variables within the UPMC data systems aggregated in its Clinical Data Warehouse. We conducted Social Security Administration Death Master File queries for vital status.^11^

### Exposure

Both mAb were provided as per their EUAs and were provided intravenously for all patients in this report. Patients who received casirivimab and imdevimab via subcutaneous injection were excluded from the analysis.

### Propensity-matched observational study of mAb treatment versus no mAb treatment

#### Patient cohort

The study population was derived from patients who received mAb treatment at UPMC infusion centers from July 14, 2021 to September 29, 2021. We derived a comparator group from the same at-risk population by identifying non-hospitalized patients with a positive polymerase chain reaction or antigen test for SARS-CoV-2 during the same time period who were eligible for mAb treatment based on EUA criteria but were not treated. For treated patients, the follow-up period began the day of treatment. For comparator patients, the follow-up period began one day after their SARS-CoV-2 test result date, which corresponded to the earliest time from test positivity to mAb treatment for treated patients. We conducted the analysis on patients treated at infusion centers, as it was not possible to identify a comparator group for patients treated in EDs.

#### Outcomes

The primary outcome was hospitalization or death by 28 days. Secondary outcomes included the 28-day rate of hospitalization, ICU admission, mechanical ventilation, and mortality.

### Statistical analysis

We performed three analyses. We used propensity matching to compared mAb-eligible patients to, i) all mAb treated patients, ii.) casirivimab and imdevimab treated patients, and iii.) sotrovimab treated patients, respectively. Propensity scores were derived from multivariable logistic regression models fit from variables with mAb treatment as the response variable and forward stepwise selection of pre-treatment explanatory variables at p < 0.15. We included variables deemed biologically relevant (e.g., age) prior to stepwise selection. We used a matching ratio (treated/untreated) of 1:2 with a maximum propensity score probability difference of 0.01. We compared characteristics of treated versus nontreated patients using Student t tests for continuous variables and χ 2 tests for categorical variables, prior to and after matching by propensity score. The primary outcome analysis consisted of the 1:2 matched comparisons of treated patients to non-treated control patients, with study outcomes expressed as an effect estimate of treatment (risk ratio, 95% confidence interval). Because of the similarity of casirivimab and imdevimab and sotrovimab treated patients (including nearly identical distributions of propensity scores), and to insure the same reference group (untreated patients) for comparison, the full set of matched controls was used in all analyses.

### Comparative effectiveness trial of casirivimab and imdevimab versus sotrovimab

The Bayesian adaptive platform trial provided the mAb therapeutic interchange via random allocation. The UPMC Quality Improvement Committee approved the study, including the random therapeutic interchange. The University of Pittsburgh IRB considered the randomized therapeutic interchange to be quality improvement and approved the additional data collection and analyses. A custom application built into the EMR linked local inventory to patient encounters and provided a random mAb allocation at time of mAb referral. Patients provided verbal consent to receive mAb therapy as part of routine care within the EUA. The prescriber and/or patient could request a specific mAb if desired.

### Patient cohort

The primary analysis population was the “as-infused” population of all patients allocated to sotrovimab or casirivimab and imdevimab, and who received mAb treatment at UPMC infusion centers and EDs during the study period. As all arms included mAb, there was no anticipated relationship between lack of infusion and the assigned arm. Due to episodic mAb shortages, some patients were treated at sites with only one available mAb at the time of treatment. We included these patients in the primary analysis as prescribing physicians and patients were blinded to drug availability at time of randomization and patients could be treated at a site different than the randomization site, pending scheduling availability.

### Outcomes

The primary outcome was hospital-free days up to day 28 after mAb treatment. This outcome is an ordinal endpoint with death as the worst outcome (labeled -1), then the length of time alive and free of hospitalization, such that the best outcome would be 28 hospital-free days. If a patient had intervening days free of hospital and was then re-hospitalized, the patient was given credit for the intervening days as free of the hospital. The secondary outcome was mortality at 28 days. We evaluated outcomes stratified by infusion location (ED versus infusion center), and the frequency of adverse events. We assessed SARS-CoV-2 variant prevalence in a random subset of enrolled patients and in our Pennsylvania catchment using Global Initiative on Sharing All Influenza Data (GISAID).^7^

### Statistical Analysis

The trial statistical analysis plan was written by blinded investigators prior to data lock and analysis. The platform is designed to continuously evaluate multiple mAb, with randomization continuing until pre-determined statistical thresholds are met. The trial launched with equal allocation randomization and planned interim analyses for adaptive randomization where mAb performing better would be given higher randomization probabilities. The mAb arm at the first adaptive analysis with the largest sample size was specified as the referent arm, as there was no non-mAb control and all patients received active treatment. Methods and results are reported as per the CONSORT Pragmatic Extension checklist (**Supplement 2**).^12^ An unblinded statistical analysis committee conducted interim and final analyses with R version 4.0.5 using the RStan package version 2.21.0 (R Foundation, Vienna, Austria) and reported results to the UPMC Chief Medical Officer who functioned in a data and safety monitor role for the study.

The primary model was a Bayesian cumulative logistic model that adjusted for treatment location (infusion center or ED), age (<30, 30-39, 40-49, 50-59, 60-69, 70-79, and ≥80 years), sex, and time (2-week epochs). The comparison between individual mAbs were based on the relative odds ratio between a given two arms for the ordinal primary outcome. An odds ratio for an arm to a comparator greater than 1 implies improved outcomes on the ordinal scale. A sliding scale with different levels of equivalence bounds was pre-defined. Equivalence between two arms was defined as a 95% posterior probability that the odds ratio is within a given odds ratio bound. Inferiority of one arm compared to another was defined as a 99% posterior probability of an odds ratio less than 1.

### Secondary analyses

We conducted a sensitivity analysis that excluded patients who received a mAb infusion at a site with only one available mAb treatment option in their “as infused” group. We conducted *a priori* subgroup analyses by vaccine status (full, partial, unvaccinated, unknown), symptom onset (>5 days, ≤5 days), and location (infusion center, ED).

### Decision to publish interim results

Due to the Delta variant public health crisis, we unblinded and analyzed patients allocated through September 29, 2021, prior to having knowingly met any pre-specified statistical threshold.

## Results

### Propensity-matched observational study of mAb treatment versus no mAb treatment

The mean age (SD) of the 1,028 treated patients was 53 (16) years versus 49 (21) years for the 5,171 patients who did not receive mAb (p < .001). Treated patients were less likely than no mAb patients to be of Black race, had lower mean Charlson Comorbidity Index scores, and more likely to have greater body mass index. After propensity score matching, treated and non-mAb treated patients were similar for all variables included in the propensity score model, the distribution of propensity scores, and variables not included in the model (**eFigure 2, eTable 1**).

Of the 1,023 propensity-matched patients who received mAb at an infusion center, 35 (3%) were hospitalized or died, and of the 2,046 propensity-matched patients who did not receive mAb, 174 (8.5%) were hospitalized or died. The mortality rate was 2.9% (n=60). In the propensity matched analysis, patients receiving mAb had lower adjusted risk of hospitalization or death (risk ratio 0.40, 95% CI: 0.28– 0.57), compared to no mAb treatment. Of the 712 propensity-matched patients who received casirivimab and imdevimab, 19 (2.7%) were hospitalized and one died (0.1%), with lower adjusted risk of hospitalization or death (risk ratio 0.31, 95% CI: 0.20–0.50), compared to no mAb treatment. Of the 311 propensity-matched patients who received sotrovimab, 16 (5.1%) were hospitalized and none died, yielding lower adjusted risk of hospitalization or death (risk ratio 0.60, 95% CI: 0.37–1.00), compared to no mAb treatment. Unmatched analyses had similar results (**eTable 2**).

### Comparative effectiveness trial of casirivimab and imdevimab versus sotrovimab

Among 4,530 referred and allocated patients, 3,558 (79%) were infused (casirivimab and imdevimab, n=2454, sotrovimab, n=1104, **Figure 1**). The mean age across groups was 54 years, half were female (54%), 12% were Black, and the most common risk factors were body mass index > 25, age ≥ 65 years, and hypertension. Mean duration of symptom onset to infusion was 5.9 days. Of those allocated to a mAb but excluded (N=972), most were not infused due to declining treatment or inability to contact (N=437, 45%), undetermined reasons (N=183, 19%), or were scheduled but not infused (N=116, 12%). One patient requested a specific mAb different than randomized assignment. All patients (N=79) tested for variant type had Delta, consistent with Pennsylvania GSAID data for this time period which showed nearly all cases were Delta (**eFigure 1**). Baseline characteristics were similar across groups (**Table 2**).

**Table 1.** Adjusted rates of hospitalization or mortality comparing mAb treatment to no mAb treatment.

|  | Number of Events |  | 28-Day Event Rate (%) |  | Risk Ratio Estimates |  |  |
| --- | --- | --- | --- | --- | --- | --- | --- |
|  | Treated | Not Treated | Treated | Not Treated | Risk Ratio | (95% CI) | p-value |
| *Casirivimab + Imdevimab vs. Not Treated* |  |  |  |  |  |  |  |
| No. | 712 | 2,046 |  |  |  |  |  |
| Hospitalization or mortality | 19 | 174 | 2.7% | 8.5% | 0.31 | (0.20–0.50) | <.001 |
| Hospitalization | 19 | 134 | 2.7% | 6.6% | 0.41 | (0.25–0.65) | <.001 |
| Mortality | 1 | 60 | 0.1% | 2.9% | 0.05 | (0.01–0.34) | .003 |
| *Sotrovimab vs. Not Treated* |  |  |  |  |  |  |  |
| No. | 311 | 2,046 |  |  |  |  |  |
| Hospitalization or mortality | 16 | 174 | 5.1% | 8.5% | 0.60 | (0.37–1.00) | .05 |
| Hospitalization | 16 | 134 | 5.1% | 6.6% | 0.79 | (0.47–1.30) | .35 |
| Mortality | 0 | 60 | 0.0% | 2.9% | 0.0 |  |  |
| *All mAb vs. Not Treated* |  |  |  |  |  |  |  |
| No. | 1,023 | 2,046 |  |  |  |  |  |
| Hospitalization or mortality | 35 | 174 | 3.4% | 8.5% | 0.40 | (0.28–0.57) | <.001 |
| Hospitalization | 35 | 134 | 3.4% | 6.6% | 0.52 | (0.36–0.75) | <.001 |
| Mortality | 1 | 60 | 0.1% | 2.9% | 0.03 | (0.01–0.34) | <.001 |

Abbreviations: CI: confidence interval

**Table 2.** Characteristics of mAb treated patients.

|  | **Casirivimab**<br>**+ Imdevimab** | **Sotrovimab** | **Entire Cohort** |
| --- | --- | --- | --- |
| No. | 2,454 (69%) | 1,104 (31%) | 3,558 |
| *Patient Characteristics* |  |  |  |
| Age, mean (SD) | 54 (18) | 53 (18) | 54 (18) |
| Female sex, n (%) | 1,320 (54%) | 599 (54%) | 1,919 (54%) |
| Race, n (%)<sup>a</sup> |  |  |  |
| White | 2,089 (85%) | 892 (81%) | 2,981 (84%) |
| Black | 252 (10%) | 161 (15%) | 413 (12%) |
| Other | 50 (2%) | 27 (2%) | 77 (2%) |
| Unknown | 63 (3%) | 24 (2%) | 87 (2%) |
| Vaccine status, n (%) |  |  |  |
| Fully vaccinated | 632 (26%) | 300 (27%) | 932 (26%) |
| Partially vaccinated | 82 (3%) | 39 (3.5%) | 121 (4%) |
| Unvaccinated | 499 (20%) | 182 (17%) | 681 (19%) |
| Unknown | 1,241 (51%) | 583 (53%) | 1,824 (51%) |
| Body mass index, mean (SD) | 32.7 (8.0) | 32.4 (7.7) | 32.6 (7.9) |
| *Timing of randomization* |  |  |  |
| Days from randomization to infusion, mean (SD) | 0.7 (1.2) | 0.6 (1.0) | 0.7 (1.2) |
| Days from symptoms to randomization, mean (SD) | 4.5 (2.0) | 4.6 (2.1) | 4.5 (2.0) |
| Days from symptoms to infusion, mean (SD) | 6.0 (1.9) | 5.8 (2.0) | 5.9 (1.9) |
| *mAb Treatment Sites* |  |  |  |
| Location, n (%) |  |  |  |
| Infusion center | 1,055 (43%) | 456 (41%) | 1,511 (43%) |
| Emergency department | 1,399 (57%) | 648 (59%) | 2,047 (58%) |
| *mAb Treatment Eligibility* |  |  |  |
| Qualifying EUA criteria, n (%) |  |  |  |
| Age ≥ 65 years | 772 (32%) | 311 (28%) | 1,083 (30%) |
| Body mass index >25 | 1,453 (59%) | 639 (58%) | 2,092 (59%) |
| Chronic kidney disease | 120 (4.9%) | 46 (4.2%) | 166 (4.7%) |
| Diabetes | 281 (11%) | 153 (14%) | 434 (12%) |
| Down syndrome | 1 (<0.1%) | 2 (0.2%) | 3 (0.1%) |
| Current or former smoker | 359 (15%) | 178 (16%) | 537 (15%) |
| Current or history of substance abuse | 26 (1.1%) | 9 (0.8%) | 35 (1.0%) |
| Immunosuppressive disease or treatment<sup>b</sup> | 383 (16%) | 169 (15%) | 552 (16%) |
| Sickle cell disease | 4 (0.2%) | 0 (0%) | 4 (0.1%) |
| Cardiovascular disease | 378 (15%) | 181 (16%) | 559 (16%) |
| Hypertension | 732 (30%) | 341 (31%) | 1,073 (30%) |
| Respiratory condition | 536 (22%) | 243 (22%) | 779 (22%) |
| Age 12–17 years with qualifying criterion | 26 (1.1%) | 6 (0.5%) | 32 (0.9%) |

<sup>a</sup>Race was reported by the patients<sup>b</sup>Immunosuppressive disease or treatment was defined as a history of HIV, cancer, transplant (solid organ, stem cell, bone marrow), chemotherapy treatment, lupus, rheumatoid arthritis, or liver disease*Abbreviations:* SD: standard deviation; EUA: emergency use authorization

### Primary outcome

The median hospital-free days were 28 (IQR 28–28) (**Table 3, Figure 2)**. Relative to the casirivimab and imdevimab group, the posterior median adjusted odds ratios from the primary model was 0.88 (95% credible interval, 0.70–1.11) for the sotrovimab group. This odds ratio resulted in an 86% probability of inferiority for sotrovimab versus casirivimab and imdevimab. The probability of equivalence between sotrovimab and casirivimab and imdevimab at the first pre-specified bound was 79%. No comparison met prespecified criteria for statistical inferiority or equivalence.

**Table 3.** Primary Outcomes from comparative effectiveness trial.

|  | **Casirivimab**<br>**+ Imdevimab** | **Sotrovimab** |
| --- | --- | --- |
| No. | 2,454 | 1,104 |
| *Primary Outcome* |  |  |
| Hospital-free days, median (IQR) | 28 (28–28) | 28 (28–28) |
| Patients with 28 hospital-free days, n (%) | 2,163 (88.1%) | 964 (87.3%) |
| Subcomponents of hospital-free days |  |  |
| Deaths, n (%) | 12 (0.5%) | 7 (0.6%) |
| Hospital length of stay among hospitalized patients, median days (IQR) | 4 (2–8) | 4 (2–8) |
| Hospitalized, n (%) | 291 (11.9%) | 140 (12.7%) |
| After mAb infusion in emergency department | 262 of 1,399 (18.7%) | 113 of 648 (17.4%) |
| After mAb infusion in infusion center | 29 of 1,055 (2.7%) | 27 of 456 (5.9%) |
| *Primary Analysis* |  |  |
| Adjusted odds ratio |  |  |
| Mean (SD) | 0.89 (0.11) | 1 [Reference] |
| Median (95% credible interval) | 0.88 (0.70–1.11) | 1 [Reference] |
| Probability of inferiority to casirivimab + imdevimab |  | 86% |
| Probability of inferiority to sotrovimab | 14% |  |
| Probability of equivalence within an odds ratio bound of, % |  |  |
| 0.25 |  | 79% |
| 0.20 |  | 67% |
| 0.15 |  | 53% |
| 0.10 |  | 36% |
| 0.05 |  | 19% |

*Abbreviations:* IQR: interquartile range; mAb: monoclonal antibodies; SD: standard deviation

**Figure 1.**
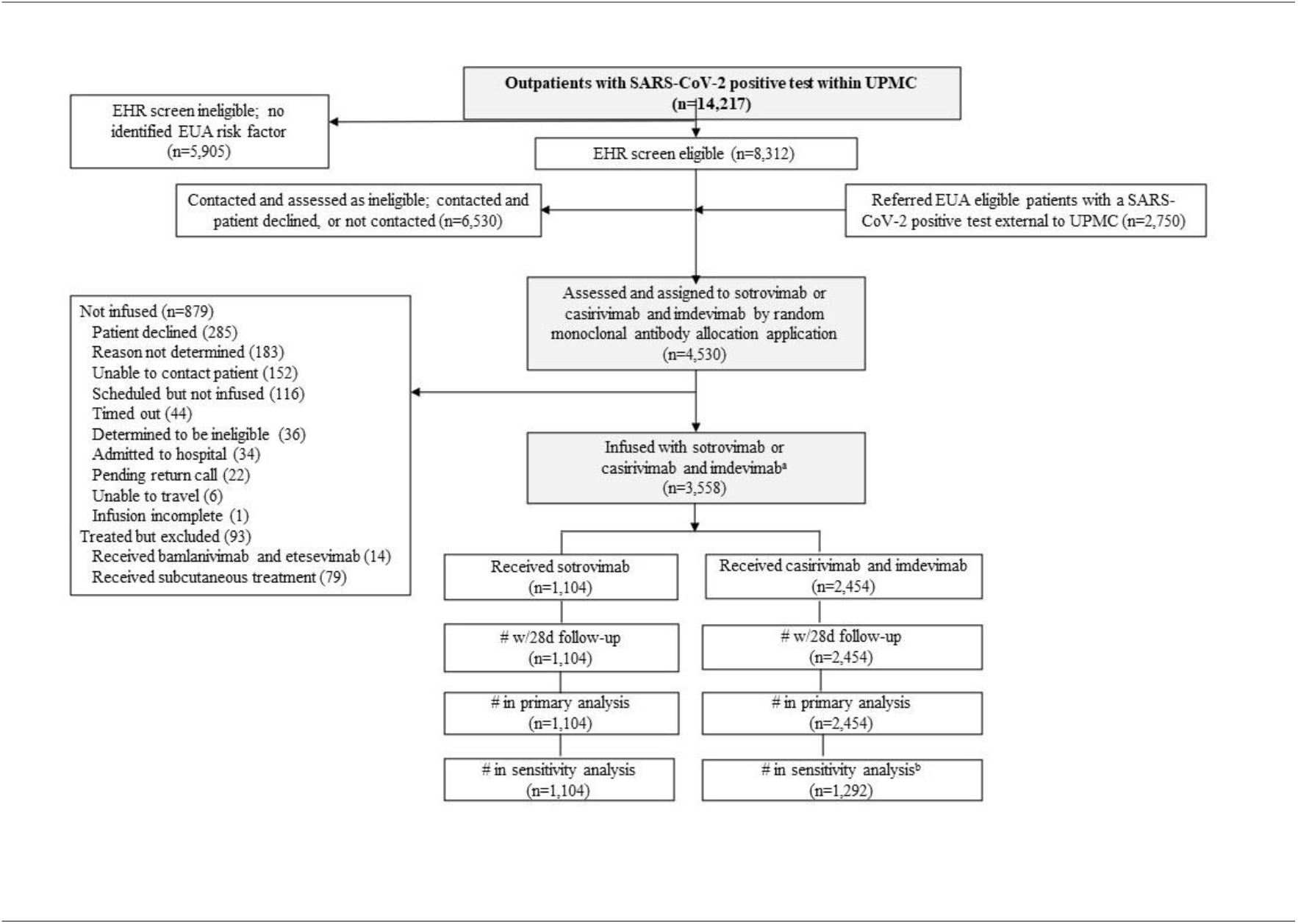
CONSORT Diagram.

**Figure 2.**
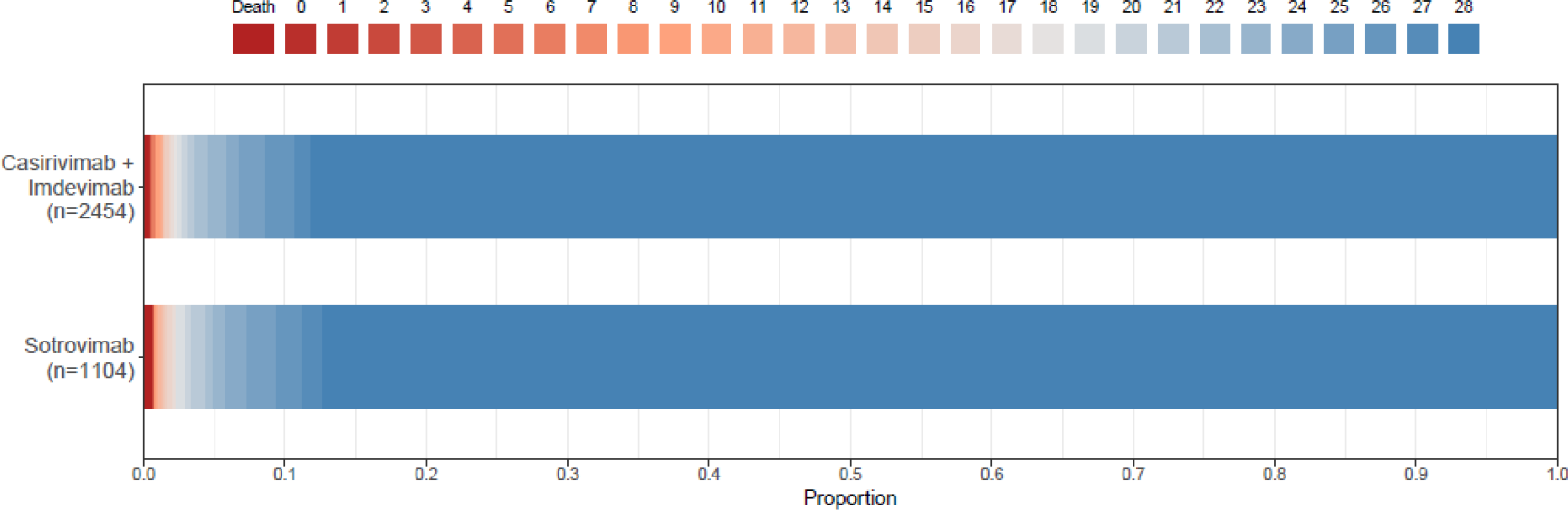
Hospital-Free Days to Day 28 in comparative effectiveness trial. Primary outcome is displayed as horizontally stacked proportions by monoclonal antibody type. Red represents worse values and blue represents better values. The median adjusted odds ratio from the primary analysis, using a Bayesian cumulative logistic model, was 0.88 (95% credible interval, 0.70–1.11) for sotrovimab versus casirivimab and imdevimab. This odds ratio yielded 86% probability of inferiority for sotrovimab versus casirivimab and imdevimab, and 79% probability of equivalence between the two at the first pre-specified bound.

### Secondary outcomes

The 28-day mortality rates were 0.5% (12/2,454) and 0.6% (7/1,104), and hospitalization rates were 11.9% (291/2,454) and 12.7% (140/1,104), in the casirivimab and imdevimab, and sotrovimab groups, respectively (**Table 3**). For patients receiving mAb in an ED, rates of hospitalization after treatment were 18.7% (262/1,399) for casirivimab and imdevimab and 17.4% (113/648) for sotrovimab. For patients receiving mAb in an infusion center, rates of hospitalization after treatment were 2.7% (29 of 1,055) for casirivimab and imdevimab and 5.9% (27 of 456) for sotrovimab.

### Sensitivity and subgroup analyses

We performed multiple sensitivity analyses to determine the robustness of our results. First, we excluded patients (n=1,162) who received casirivimab and imdevimab at a location where only casirivimab and imdevimab was available and found similar results (81% probability of sotrovimab inferiority to casirivimab and imdevimab, 79% equivalence). No subgroup analysis except location met the pre-specified thresholds for statistical inferiority or equivalence. Among the 1,511 patients treated at an infusion center, the median odds ratio of sotrovimab compared to casirivimab and imdevimab was 0.48 (95% credible interval, 0.28–0.84), resulting in a 99.5% probability of inferiority.

### Adverse events

Among the 2,454 patients treated with casirivimab and imdevimab, adverse events (AE) and serious adverse events (SAE) were rare, reported in 17 (0.5%) and 7 (0.2%) of patients, respectively. Similarly, for the 1,104 sotrovimab treated patients, respective rates were 0.5% (6 patients) and 0.4% (4 patients). The most commonly reported AE were flushing, itching, breathing difficulties, and chest tightness/pain.

## Discussion

During the COVID-19 Delta variant surge, we found that casirivimab-imdevimab and sotrovimab were each associated with a reduced risk-adjusted hospitalization and death among patients with mild to moderate COVID-19 compared to no treatment. In a comparative effectiveness randomized trial, no primary analysis met a prespecified trigger for conclusions of inferiority or equivalence, although the subgroup analysis of patients treated in an infusion center found superiority of casirivimab-imdevimab over sotrovimab.

The Delta variant of SARS-CoV-2 was observed late 2020, and by July 2021 was the dominant strain worldwide. Yet, early evidence that demonstrated the efficacy of mAb were conducted prior to the emergence of Delta. Our data extend earlier work to show that mAbs are associated with improved outcomes in Delta in a cohort with a robust sample size and methods to adjust for treatment selection and confounding. Future work will determine effectiveness versus the Omicron variant.

We found no difference when comparing the effectiveness of mAb according to predefined statistical thresholds. This finding could be due to true absence of a difference or from insufficient power to detect a difference due to our decision to evaluate early because of pandemic emergency. In an *a priori* subgroup, we did observe a 99.5% probability of inferiority in patients treated at an infusion center. Although not a primary analysis, this finding, if replicated, suggests that at least for patients with the Delta variant treated in an infusion center, casirivimab-imdevimab may be preferred. Patients presenting to an ED may have a greater illness severity compared to patients presenting to a scheduled appointment at an infusion center, and their illness severity may be the main determinant of their subsequent course, not mAb type received.

There are many limitations to these studies. First, the observational analysis of mAb treatment versus no mAb treatment may be subject to residual or unmeasured confounding. Second, we evaluated comparative effectiveness trial results early due to the Delta variant crisis and the primary analysis was inconclusive. Third, this report is restricted to the patients in the catchment of UPMC, limiting generalizability. Finally, we were not powered to determine treatment effects by symptom onset or vaccine status.

## Conclusion

In non-hospitalized patients with mild to moderate COVID-19 due to the Delta variant, casirivimab and imdevimab, and sotrovimab were both associated with reduced risk of hospitalization or death. The comparative effectiveness of mAbs appeared similar, though prespecified criteria for statistical inferiority or equivalence were not met.

## Data Availability

All data produced in the present study are available upon reasonable request to the authors.

## Article Information

## Acknowledgements

The authors thank the clinical staff of the UPMC monoclonal antibody infusion centers as well as the support and administrative staff behind this effort, including but not limited to: Michelle Adam, Jodi Ayers, Ashley Beyerl, Trudy Bloomquist, Tina Borneman, Mikaela Bortot, Jonya Brooks, James Cable, Sherry Casali, Jeana Colella, Jennifer Dueweke, Jesse Duff, Janice Dunsavage, Jessica Fesz, Kathleen Flinn, Daniel Gessel, Amy Helmuth, Erik Hernandez, Larry Hruska, Allison Hydzik, Le Ann Kaltenbaugh, LuAnn King, Jim Krosse, Sheila Kruman, Amy Lukanski, Hilary Maskiewicz, Debra Masser, Katelyn Mayak, Rebecca Medva, Theresa Murillo, Melanie Pierce, Teressa Polcha, Kevin Pruznak, Debra Rogers, Rozalyn Russell, Sarah Sakaluk, Heather Schaeffer, Robert Shulik, Libby Shumaker, Susan Spencer, Ashley Steiner, Betsy Tedesco, Ken Trimmer, Jennifer Zabala, and their entire teams.

## Affiliations of Authors/members of the writing committee

We acknowledge staff at UPMC Clinical Analytics, the UPMC Wolff Center, and Biostatistics and Data Management Core at the CRISMA Center in the Department of Critical Care Medicine at the University of Pittsburgh for curating and managing the data.

## Author contributions

Dr. Huang and Seymour take full responsibility for the integrity of the data and the accuracy of the data analysis.

*Study concept and design:* Huang, McCreary, Kip, Seymour

*Acquisition of data:* Huang, McCreary, Collins, Kip, Seymour

*Interpretation of data:* All authors

*Drafting of the manuscript:* Huang, Seymour

*Critical revision of the manuscript for important intellectual content:* All authors

*Study supervision:* Huang, Seymour

## Supplementary Materials

### Supplement 1 - Online Supplemental Materials

**eFigure 1.**
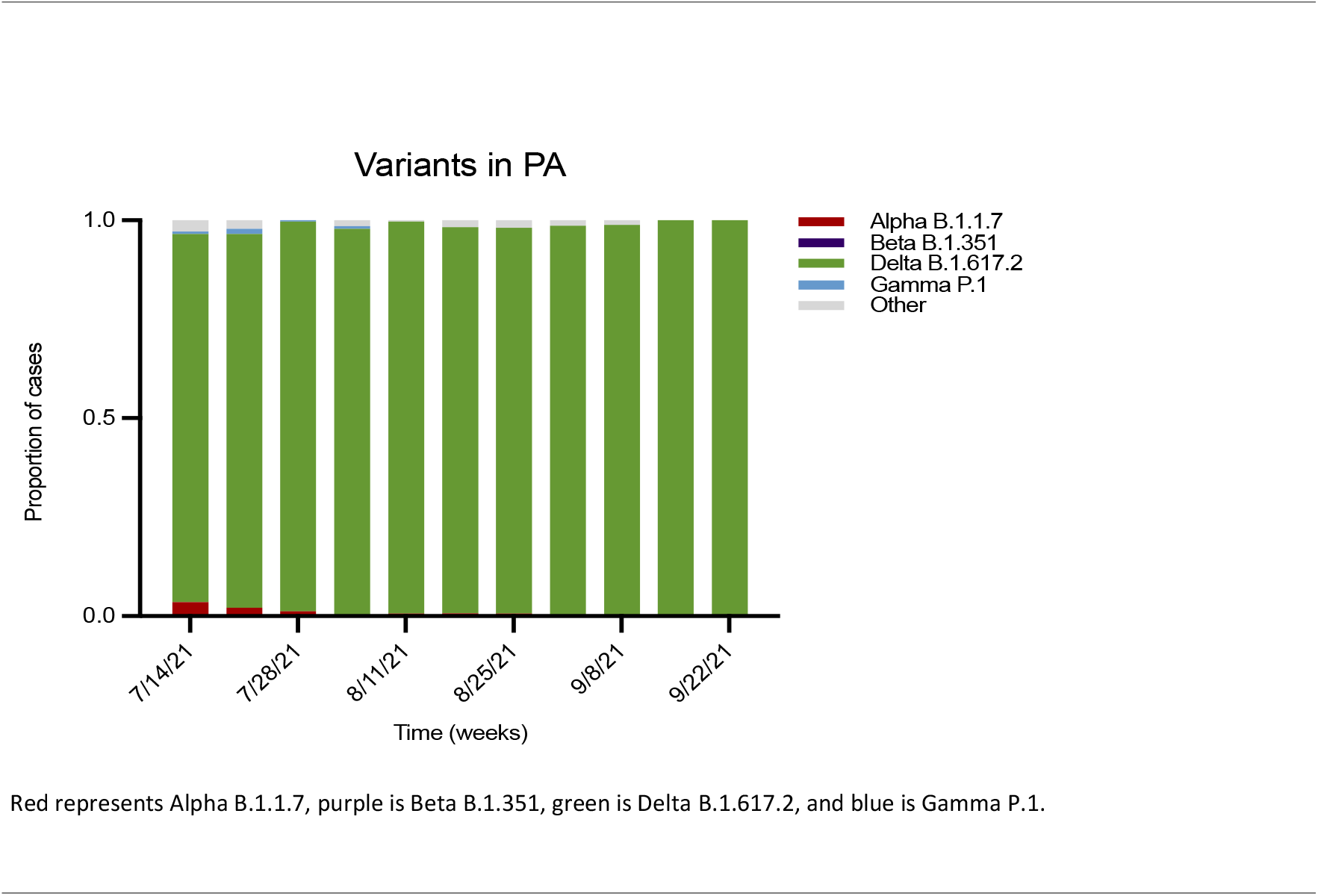
SARS-CoV-2 Variants of Concern Proportion in Pennsylvania During the Study(July 14 – September 22, 2021).

**eFigure 2.**
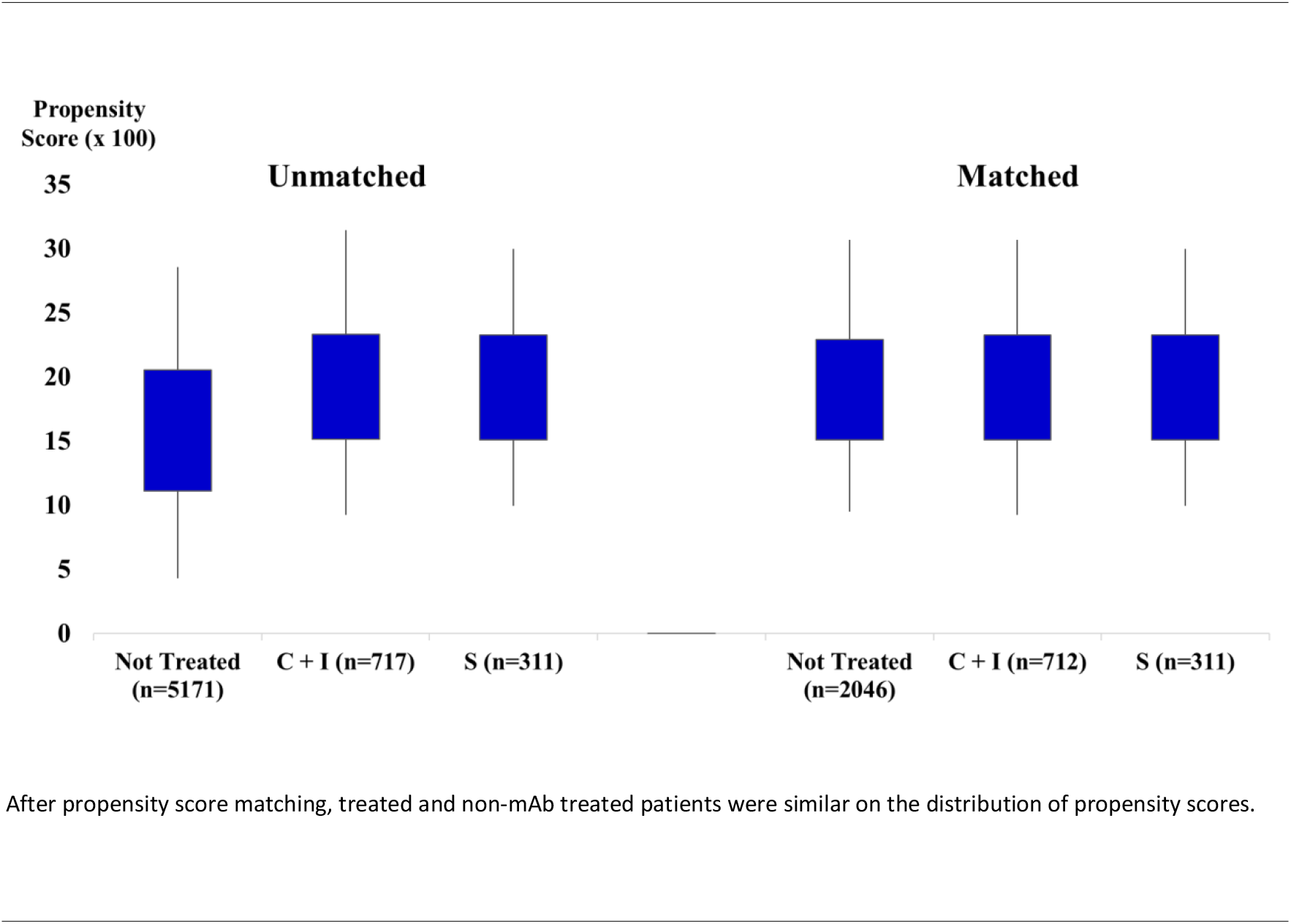
Propensity Scores for Unmatched and Matched Patients.

**eTable 1.** Comparison of Characteristics of Unmatched and Propensity Matched Patients.

|  | <u>Unmatched</u> |  |  | <u>Propensity Matched</u> |  |  |
| --- | --- | --- | --- | --- | --- | --- |
|  | Treated | Not Treated | p-value | Treated | Not Treated | p-value |
| No. | 1,028 | 5,171 |  | 1,023 | 2,046 |  |
| *Patient Characteristics* |  |  |  |  |  |  |
| Age, mean (SD) | 53.2 (16.4) | 49.6 (21.4) | <.001 | 53.2 (16.4) | 52.8 (19.5) | .50 |
| Female gender, (No.), % | (572) 55.6% | (3013) 58.3% | .12 | (569) 55.6% | (1157) 56.6% | .62 |
| Black race, (No.), % | (61) 5.9% | (590) 11.4% | <.001 | (61) 6.0% | (112) 5.5% | .58 |
| Body mass index, mean (SD) | 32.5 (7.4) | 31.8 (7.5) | .008 | 32.5 (7.4) | 32.6 (7.8) | .60 |
| *Medical History* |  |  |  |  |  |  |
| History of cancer, (No.), % | (46) 4.5% | (597) 11.6% | <.001 | (42) 4.1% | (105) 5.1% | .21 |
| History of congestive heart failure, (No.), % | (47) 4.6% | (399) 7.7% | <.001 | (46) 4.5% | (93) 4.6% | .95 |
| History of obstructive sleep apnea, (No.), % | (198) 19.3% | (743) 14.4% | <.001 | (195) 19.1% | (338) 16.5% | .08 |
| Charlson Comorbidity Index Score, mean (SD) | 0.7 (0.9) | 0.9 (1.2) | <.001 | 0.7 (0.9) | 0.7 (0.9) | .57 |
| History of rheumatoid arthritis, (No.), % | (38) 3.7% | (104) 2.0% | .001 | (34) 3.3% | (61) 3.0% | .61 |
| History of atrial fibrillation, (No.), % | (46) 4.5% | (307) 5.9% | .06 | (46) 4.5% | (90) 4.4% | .90 |
| History of valvular heart disease, (No.), % | (73) 7.1% | (287) 5.6% | .05 | (72) 7.0% | (138) 6.7% | .76 |
| History of viral hepatitis, (No.), % | (7) 0.7% | (79) 1.5% | .03 | (7) 0.7% | (16) 0.8% | .77 |
| Antidepressants, (No.), % | (335) 32.6% | (1497) 29.0% | .02 | (330) 32.3% | (674) 32.9% | .70 |
| History of asthma, (No.), % | (312) 30.4% | (1851) 35.8% | <.001 | (311) 30.4% | (593) 29.0% | .42 |
| History of stroke, (No.), % | (45) 4.4% | (305) 5.9% | .05 | (45) 4.4% | (91) 4.5% | .95 |
| History of allergic rhinitis, (No.), % | (146) 14.2% | (658) 12.7% | .20 | (145) 14.2% | (268) 13.1% | .41 |
| Alpha blockers, (No.), % | (11) 1.1% | (78) 1.5% | .28 | (11) 1.1% | (20) 1.0% | .80 |
| *Variables Not Included in Propensity Model* |  |  |  |  |  |  |
| ACE Inhibitors, (No.), % | (144) 14.0% | (759) 14.7% | .58 | (144) 14.1% | (303) 14.8% | .59 |
| Angiotensin II receptor blocker, (No.), % | (104) 10.1% | (450) 8.7% | .15 | (102) 10.0% | (195) 9.5% | .70 |
| Beta blockers, (No.), % | (215) 20.9% | (1114) 21.5% | .65 | (212) 20.7% | (462) 22.6% | .24 |
| History of coronary artery disease, (No.), % | (104) 10.1% | (542) 10.5% | .73 | (103) 10.1% | (188) 9.2% | .43 |
| History of chronic kidney disease, (No.), % | (53) 5.2% | (336) 6.5% | .11 | (53) 5.2% | (117) 5.7% | .54 |
| History of COPD, (No.), % | (148) 14.4% | (831) 16.1% | .18 | (146) 14.3% | (272) 13.3% | .46 |
| Corticosteroids, (No.), % | (348) 33.9% | (1639) 31.7% | .18 | (345) 33.7% | (653) 31.9% | .31 |
| History of diabetes, (No.), % | (167) 16.3% | (883) 17.1% | .52 | (166) 16.2% | (331) 16.2% | .97 |
| History of dyspnea, (No.), % | (51) 5.0% | (295) 5.7% | .34 | (51) 5.0% | (104) 5.1% | .91 |
| History of fatty liver disease, (No.), % | (34) 3.3% | (143) 2.8% | .34 | (34) 3.3% | (59) 2.9% | .50 |
| History of hypertension, (No.), % | (426) 41.4% | (2070) 40.0% | .40 | (423) 41.4% | (861) 42.1% | .70 |
| History of pulmonary hypertension, (No.), % | (8) 0.8% | (102) 2.0% | .008 | (8) 0.8% | (30) 1.5% | .11 |
| Statins, (No.), % | (320) 31.1% | (1485) 28.7% | .12 | (318) 31.1% | (630) 30.8% | .87 |

**eTable 2.**
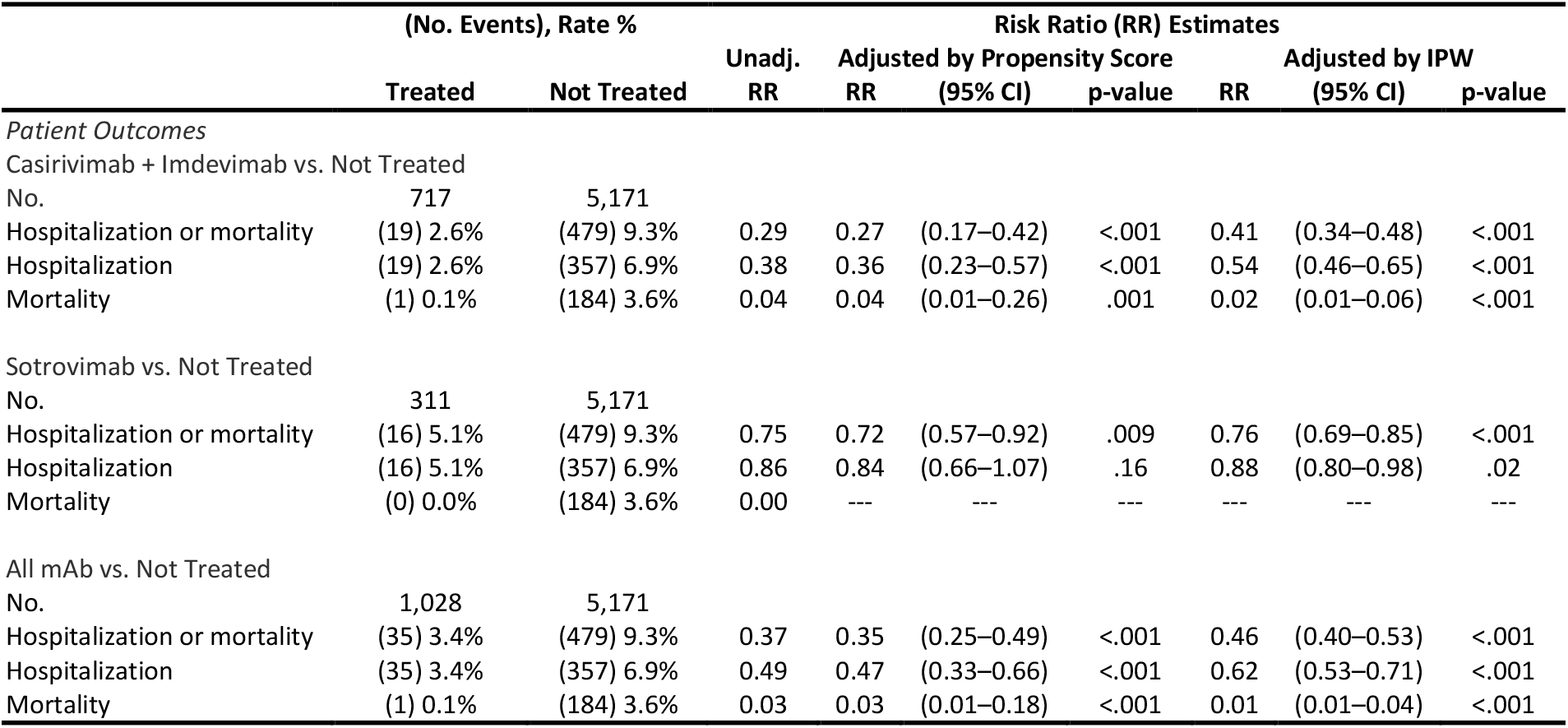
Primary and Secondary Outcomes in an Unmatched Cohort of Patients Receiving Monoclonal Antibody Treatment and an At-Risk Population of Patients Not Receiving Monoclonal Antibody Treatment.

### Supplement 2 - CONSORT Pragmatic Trials Checklist

| Section | Item | Standard CONSORT Description | Pragmatic Trials Extension | Page |
| --- | --- | --- | --- | --- |
| Title and abstract | 1 | How participants were allocated to interventions (e.g., “random allocation,” “randomized,” or “randomly assigned”) |  | Title page 1 & Abstract pages 4-5 |
| **Introduction** |  |  |  |  |
| Background | 2 | Scientific background and explanation of rationale | Describe the health or health service problem that the intervention is intended to address and other interventions that may commonly be aimed at this problem | 6 |
| **Methods** |  |  |  |  |
| Participants | 3 | Eligibility criteria for participants; settings and locations where the data were collected | Eligibility criteria should be explicitly framed to show the degree to which they include typical participants and/or, where applicable, typical providers (e.g., nurses), institutions (e.g., hospitals), communities (or localities e.g., towns) and settings of care (e.g., different healthcare financing systems) | 8-9 |
| Interventions | 4 | Precise details of the interventions intended for each group and how and when they were actually administered | Describe extra resources added to (or resources removed from) usual settings in order to implement intervention. Indicate if efforts were made to standardize the intervention or if the intervention and its delivery were allowed to vary between participants, practitioners, or study sites<br>Describe the comparator in similar detail to the intervention | —<br>— |
| Objectives | 5 | Specific objectives and hypotheses |  | 6 |
| Outcomes | 6 | Clearly defined primary and secondary outcome measures and, when applicable, any methods used to enhance the quality of measurements (e.g., multiple observations, training of assessors) | Explain why the chosen outcomes and, when relevant, the length of follow-up is considered important to those who will use the results of the trial | 7-8 |
| Sample size | 7 | How sample size was determined; explanation of any interim analyses and stopping rules when applicable | If calculated using the smallest difference considered important by the target decision maker audience (the minimally important difference) then report where this difference was obtained | — |
| Randomization—sequence generation | 8 | Method used to generate the random allocation sequence, including details of any restriction (e.g., blocking, stratification) |  | — |
| Randomization—allocation concealment | 9 | Method used to implement the random allocation sequence (e.g., numbered containers or central telephone), clarifying whether the sequence was concealed until interventions were assigned |  | 11-12 |
| Randomization—implementation | 10 | Who generated the allocation sequence, who enrolled participants, and who assigned participants to their groups |  | — |
| Blinding (masking) | 11 | Whether participants, those administering the interventions, and those assessing the outcomes were blinded to group assignment | If blinding was not done, or was not possible, explain why | 11 |
| Statistical methods | 12 | Statistical methods used to compare groups for primary outcomes; methods for additional analyses, such as subgroup analyses and adjusted analyses |  | 11-12 |
| **Results** |  |  |  |  |
| Participant flow | 13 | Flow of participants through each stage (a diagram is strongly recommended)—specifically, for each group, report the numbers of participants randomly assigned, receiving intended treatment, completing the study protocol, and analyzed for the primary outcome; describe deviations from planned study protocol, together with reasons | The number of participants or units approached to take part in the trial, the number which were eligible, and reasons for non-participation should be reported | 11 |
| Recruitment | 14 | Dates defining the periods of recruitment and follow-up |  | — |
| Baseline data | 15 | Baseline demographic and clinical characteristics of each group |  | 13 |
| Numbers analyzed | 16 | Number of participants (denominator) in each group included in each analysis and whether analysis was by “intention-to-treat;” state the results in absolute numbers when feasible (e.g., 10/20, not 50%) |  | 13 |
| Outcomes and estimation | 17 | For each primary and secondary outcome, a summary of results for each group and the estimated effect size and its precision (e.g., 95% CI) |  | 14-15 |
| Ancillary analyses | 18 | Address multiplicity by reporting any other analyses performed, including subgroup analyses and adjusted analyses, indicating which are prespecified and which are exploratory |  | 15 |
| Adverse events | 19 | All-important adverse events or side effects in each intervention group |  | 15 |
| **Discussion** |  |  |  |  |
| Interpretation | 20 | Interpretation of the results, considering study hypotheses, sources of potential bias or imprecision, and the dangers associated with multiplicity of analyses and outcomes |  | 16-17 |
| Generalizability | 21 | Generalizability (external validity) of the trial findings | Describe key aspects of the setting which determined the trial results. Discuss possible differences in other settings where clinical traditions, health service organization, staffing, or resources may vary from those of the trial | 16-18 |
| Overall evidence | 22 | General interpretation of the results in the context of current evidence |  | 18 |

